# NCBoost v2: a classifier for non-coding variants in Mendelian diseases

**DOI:** 10.1101/2025.09.18.25336072

**Authors:** Barthélémy Caron, Antonio Rausell

**Affiliations:** Université Paris Cité, INSERM UMR1163, Imagine Institute, Clinical Bioinformatics Laboratory, Paris, F-75006, France; AP-HP, Necker Hospital for Sick Children, Fédération de Génétique et Médecine Génomique, Service de Médecine Génomique des Maladies Rares, Paris, F-75015, France

## Abstract

**Motivation:** The current diagnostic rate of rare diseases through whole-genome sequencing has stabilized at around 30% on average, highlighting the need for improved computational scores to identify pathogenic variants. In 2019, we developed NCBoost, a supervised-learning approach that mined a comprehensive set of sequence constraint features and proved particularly well suited to identifying high-effect pathogenic non-coding variants in genetic diseases. Since its first release, the substantial increase in the number of variants available for training, as well as the enhanced capacity to detect purifying selection signals from large-scale genome sequencing projects, motivated an update of NCBoost.

**Results:** We implemented NCBoost v2, a pathogenicity score for non-coding single-nucleotide variants, trained on the largest set of curated pathogenic variants in monogenic Mendelian diseases available to date. It leverages conservation features computed from recent large-scale genomic consortia such as Zoonomia and gnomAD, and incorporates recent splice-altering predictive scores. NCBoost v2 outperformed alternative state-of-the-art methods in a variety of scenarios, providing more consistent scores across non-coding genomic regions and fine-tuning the scoring of pathogenic splice-altering variants in Mendelian disease genes.

**Availability:** NCBoost v2 software is implemented in Python 3.10 and is freely available under the GNU General Public License Version 3 at https://doi.org/10.5281/zenodo.16029049 and https://github.com/RausellLab/NCBoost-2, together with precomputed scores for the human genome assembly GRCh38.

## 1 Introduction

Rare diseases affect approximately 5–8% of the global population (Kernohan and Boycott 2024), with more than 7,000 distinct disorders described to date (*OMIM - (OMIM*.*ORG)*). Whole genome sequencing has become a first-line clinical diagnosis test for investigating their genetic basis, which is estimated to underlie 80% of rare diseases. Its adoption in national genomic medicine plans has demonstrated its added value in improving diagnostic yield, including the identification of novel causal variants in non-coding genomic regions, among other factors (PFMG contributors, 2025). Yet, WGS yields a molecular diagnosis in only 30–50% of suspected Mendelian cases, depending on disease type (Kaur *et al*. 2024). The important fraction of unresolved patients highlights the need for improved methods to prioritize genetic variants, particularly in non-coding regions, which make up 98% of the human genome. A number of computational scoring methods have contributed to the identification of causal non-coding variants (Fu *et al*. 2014; Kircher *et al*. 2014; Zhou and Troyanskaya 2015; Ionita-Laza *et al*. 2016; Smedley *et al*. 2016), including our NCBoost method (Caron, Luo and Rausell 2019). NCBoost is a supervised learning approach trained to discriminate pathogenic from benign non-coding variants by leveraging a comprehensive set of interspecies conservation features, as well as recent and ongoing sequence constraint signals in the human lineage. NCBoost proved to be particularly well-suited for the assessment of monogenic genetic diseases, which are often driven by high-effect causal variants subject to purifying selection. NCBoost was referred to in the recommendations for clinical interpretation of variants found in non-coding regions of the genome published in Ellingford et al., (2022). Since its first release in 2019, the widespread adoption of WGS has translated in a significant increase in the number of reported causal variants in non-coding regions in reference databases for rare disease patients such ClinVar (Landrum *et al*. 2014) and HGMD professional (Stenson *et al*. 2020). In addition, recent large-scale sequencing projects in vertebrates, mammals, primates as well as of the general population, are permitting the identification of purifying selection signals in genomic sequences with enhanced resolution (Zoonomia Consortium et al. 2020, Chen et al. 2024). Furthermore, novel computational methods have greatly improved the identification of splice-altering variants (Jaganathan *et al*. 2019, Cheng *et al*. 2019, Wagner *et al*. 2023), and have been integrated within genetic diagnosis protocols (Walker *et al*. 2023). To take advantage of the previous developments, we present NCBoost v2, an updated version of the NCBoost approach on the newer human genome assembly hg38 that is trained on a significantly larger set of high-confidence pathogenic non-coding variants associated with monogenic Mendelian diseases, annotated with more comprehensive conservation features computed on recent large-scale genomic projects as well as with splice-altering scores, and thoroughly tested and validated in multiple scenarios.

## 2 Methods

NCBoost v2 is a supervised learning algorithm using extreme gradient boosting (XGBoost) and trained to discriminate non-coding pathogenic single nucleotide variants (SNVs) associated with monogenic Mendelian diseases (MMDs) from non-pathogenic SNVs. Non-coding pathogenic variants were obtained from HGMD professional (Stenson *et al*. 2020) and ClinVar (Landrum *et al*. 2014) and extensive curation steps were followed to obtain a high-confidence set of N=2,336 non-coding SNVs causing MMDs, collectively involving 764 unique genes (**Figure S1** and **Supplementary Text**). This dataset is significantly larger than the analogous set available at the time of the first release of NCBoost (737 variants associated with 283 unique genes). Random common variants were sampled from dbSNP (Sherry 2001) and used as the negative training set. To avoid cross-contamination between training and downstream application sets, NCBoost v2 was trained as a bundle of 10 independent models as follows: protein-coding genes were randomly split into 10 genomic partitions, equally stratified across chromosomes, with each containing a similar number of genes with and without high-quality non-coding pathogenic variants. Each positive non-coding SNV was paired with 10 random negative SNVs from the same genomic partition and the same type of genetic region (upstream, 5’UTR, intronic, 3’UTR, downstream and intergenic). Throughout these steps, a maximum of 1 positive and 1 negative variant per gene were sampled, while maintaining a global 1:10 positive-to-negative ratio in the training set (**Supplementary Text**).

Variants were annotated with 47 features, including 15 new ones compared to the initial NCBoost version, belonging to four different feature categories (**Table S1** and **Supplementary Text**): (A) long-term interspecies sequence conservation features, including recent large-scale alignment of 241 vertebrate genomes and 43 primate genomes from the Zoonomia project (Zoonomia Consortium *et al*. 2020); (B) recent and ongoing human sequence constraint features, computed on more than 76,000 genomes from GnomAD v4.1, representing nine major subpopulations (Chen *et al*. 2024b); (C) conservation features of the associated protein-coding gene, including both interspecies and in-human sequence conservation, such as the *loeuf* score, computed on more than 730,000 individuals from GnomAD (Karczewski *et al*. 2020); and (D) sequence context features, such as GC and CpG content around the position, the type of overlapping non-coding region and the maximum SpliceAI score at the position, in order to inform the classifier about its splice-altering potential (**Supplementary Text** and **Figure S2**).

## 3 Results

NCBoost v2 was first cross-validated on each of the 10 genomic partitions, iteratively left out during the training phase, collectively adding a total of 736 high-quality non-coding pathogenic variants and the corresponding region-matched set of 7,360 random common non-coding variants (**Table 1, Figure S3** and **Supplementary Text**). NCBoost v2 achieved an area under the receiver operating characteristic (AUROC) curve of 0.93 and a precision-recall (AUPR) curve of 0.77, outperforming two alternative methods, representative of the state-of-the-art, such as ReMM (Schubach et al., 2022) and CADD (Schubach et al. 2024). NCBoost v2’s superiority was consistent across all genomic region types, independently considered. (**Table 1**). Outperformance of NCBoost v2 was also consistently observed when excluding SpliceAI from the feature set, whether considered globally or for each genomic region (**Table S2**). For the remainder of this work, NCBoost v2 refers to the framework trained with SpliceAI included as a feature.

**Table 1.** The table reports the ability of NCBoost v2 to discriminate between pathogenic and non-pathogenic non-coding variants, together with two alternative state-of-the-art methods: ReMM and CADD. The area under the receiver operating characteristic (AUROC) curve and the area under the precision-recall (AUPR) curve are reported for each method across three evaluation settings: genomic partition cross-validation, independent testing, and validation (**Methods** and **Supplementary Text**). For each setting, the global AUROC and AUPR values are reported, along with those corresponding to the different region types evaluated independently. The total number of pathogenic and non-pathogenic variants is reported for each setting, as well as broken down by region type.

|  | Region | N patho-genic | N benign | NCBoost v2 |  | ReMM |  | CADD |  |
| --- | --- | --- | --- | --- | --- | --- | --- | --- | --- |
|  |  |  |  | AUROC | AUPR | AUROC | AUPR | AUROC | AUPR |
| Genomic-partition cross-validation | **Global** | **763** | **7630** | **0.93** | **0.77** | **0.72** | **0.24** | **0.72** | **0.35** |
|  | Upstream | 59 | 590 | 0.88 | 0.55 | 0.85 | 0.48 | 0.79 | 0.46 |
|  | UTR5 | 133 | 1330 | 0.81 | 0.40 | 0.71 | 0.24 | 0.71 | 0.27 |
|  | Intronic | 475 | 4750 | 0.97 | 0.89 | 0.72 | 0.25 | 0.71 | 0.40 |
|  | UTR3 | 81 | 810 | 0.85 | 0.56 | 0.73 | 0.29 | 0.74 | 0.63 |
|  | Downstream | 5 | 50 | 0.87 | 0.58 | 0.81 | 0.45 | 0.89 | 0.43 |
|  | Intergenic | 10 | 100 | 0.77 | 0.48 | 0.79 | 0.42 | 0.69 | 0.40 |
| Testing | **Global** | **687** | **6870** | **0.88** | **0.59** | **0.79** | **0.43** | **0.78** | **0.43** |
|  | Upstream | 177 | 1770 | 0.90 | 0.67 | 0.86 | 0.57 | 0.86 | 0.58 |
|  | UTR5 | 252 | 2520 | 0.81 | 0.32 | 0.77 | 0.43 | 0.76 | 0.42 |
|  | Intronic | 156 | 1560 | 0.97 | 0.86 | 0.80 | 0.45 | 0.73 | 0.43 |
|  | UTR3 | 80 | 800 | 0.84 | 0.49 | 0.78 | 0.32 | 0.76 | 0.40 |
|  | Downstream | 9 | 90 | 0.97 | 0.85 | 0.82 | 0.67 | 0.91 | 0.74 |
|  | Intergenic | 13 | 130 | 0.89 | 0.69 | 0.84 | 0.65 | 0.81 | 0.61 |
| Validation | **Global** | **151** | **1510** | **0.96** | **0.84** | **0.71** | **0.25** | **0.74** | **0.35** |
|  | Upstream | 1 | 10 | 1.00 | 1.00 | 1.00 | 1.00 | 1.00 | 1.00 |
|  | UTR5 | 18 | 180 | 0.91 | 0.57 | 0.78 | 0.32 | 0.69 | 0.33 |
|  | Intronic | 123 | 1230 | 0.97 | 0.90 | 0.72 | 0.28 | 0.76 | 0.39 |
|  | UTR3 | 12 | 120 | 0.78 | 0.39 | 0.69 | 0.29 | 0.66 | 0.25 |
|  | Downstream | 1 | 10 | 1.00 | 1.00 | 0.40 | 0.07 | 0.60 | 0.10 |
|  | Intergenic | 3 | 30 | 0.92 | 0.75 | 0.61 | 0.41 | 0.69 | 0.56 |

Consistent with our previous observations (Caron et al., 2019), per-gene region biases were observed across common SNVs without clinical assertions, with the 5’ UTR showing the strongest shift towards higher values, suggesting that the regulatory region where a variant maps systematically biases the scores (**Figure S4** and **Supplementary Text**). However, only in the case of NCBoost v2 were the distributions of median scores per gene for common SNVs in 5’ UTRs significantly different (with less severe scores) from those of pathogenic SNVs in the 5’ UTR as well as in upstream, intronic, 3’ UTR, and downstream regions of the associated genes, while ReMM and CADD failed to overcome such bias in several of those regions (**Table S3**).

We further evaluated NCBoost v2 on an independent test set of N=687 high-confidence, manually curated non-coding pathogenic variants reported at the time of NCBoost’s first release (Caron et al., 2019), as well as in a validation set of N=151 non-coding pathogenic variants reported after January 2015 (**Supplementary Text**). NCBoost’s capacity to discriminate pathogenic from non-pathogenic variants generalized both in the test and validation sets (**Figures S5** and **S6**), was consistent across all gene regions evaluated, and systematically outperformed alternative scores (**Table 1**).

To test NCBoost v2 performance in more realistic clinical diagnosis scenarios, we simulated disease genomes by introducing within each genome of 100 healthy individuals, independently, each of the non-coding pathogenic variants from the validation set along with a random common variant mapping to the same non-coding region of the same gene (**Supplementary Text**). While the three evaluated scores ranked the non-coding pathogenic variants significantly higher than the matched common variants (one-sided paired Wilcoxon test: NCBoost v2 p-value = 1.41e-32, ReMM p-value = 9.54e-11, and CADD p-value = 1.24e-13), NCBoost v2 showed the best within-individual ranking of non-coding pathogenic variants (median = 99.93%), significantly outperforming ReMM (median=92.94, one-sided paired Wilcoxon test p-value=4.95e-25) and CADD (median=92.57%, p-value= 6.30e-28, **Figure S6**).

Finally, given the high importance of SpliceAI as a predictive feature in the NCBoost v2 model (**Figure S2**), we evaluated whether NCBoost v2 provides an added value compared to using SpliceAI as a raw score. To do this, for each protein-coding gene, we obtained the top-scoring SpliceAI non-coding position’s score along with its corresponding NCBoost v2 score. We then compared the gene rank percentiles of such scores across N=6,128 MMD genes and N=12,640 non-disease genes. While the distribution of both scores were significantly higher across MMD genes than non-disease genes (two-sided Mann-Whitney-Wilcoxon rank-sum test p-value = 4.3e-23 and 1.2e-102, for SpliceAI and NCBoost v2, respectively), NCBoost v2 provided higher ranks (i.e. more pathogenic) to MMDGs splice-altering variants than SpliceAI (two-sided Mann-Whitney-Wilcoxon rank-sum test p-value = 1.3e-17, and two-sided Wilcoxon paired signed-rank test pvalue = 9.2e-08, **Figure S8** and **Supplementary Text**).

## Discussion

NCBoost v2 proved to be a valuable supervised-learning approach for identifying pathogenic non-coding variants in monogenic Mendelian diseases across diverse independent test and validation settings. NCBoost outperformed reference state-of-the-art scores recently updated for the newer human genome assembly, hg38. Its superiority was consistent across all genomic region types, considered independently. Notwithstanding, consistent with our previous observations (Caron et al., 2019), per-gene region biases were still observed for all evaluated scores. However, NCBoost v2 was the only score capable of assigning more severe scores to pathogenic than to non-pathogenic variants, independently of the gene region considered (i.e., upstream, 5’UTR, intronic, 3’UTR, and downstream regions). Finally, the incorporation of SpliceAI scores as features into the NCBoost v2 framework resulted in a score that better discriminates splice-altering variants in monogenic Mendelian disease genes compared to non-disease genes, improving the raw SpliceAI values. These results suggest that NCBoost v2’s integration of variant- and gene-level conservation features helps fine-tuning SpliceAI predictions for improved application in rare disease genomes. The NCBoost v2 score should be used in the context of more comprehensive genetic diagnostic protocols and guidelines, further evaluating the consistency between altered genomic elements and observed clinical phenotypes, among other factors in rare disease clinical assessment (Ellingford et al., 2022; Walker et al., 2023).

## Data Availability

NCBoost v2 software is implemented in Python 3.10 and is freely available under the GNU General Public License Version 3 at https://doi.org/10.5281/zenodo.16029049 and https://github.com/RausellLab/NCBoost-2, together with precomputed scores for the human genome assembly GRCh38.

## Author contributions

Conceptualization: BC & AR; Data curation: BC; Formal analysis: BC; Funding acquisition: AR; Investigation: BC & AR; Methodology: BC & AR; Project administration: BC & AR; Resources: BC & AR; Software: BC; Supervision: AR; Validation: BC & AR; Visualization: BC; Writing – original draft: BC & AR; Writing – review & editing: BC & AR.

## Acknowledgements

We thank the Bioinformatics platform and IT service of the Institute Imagine for continuous support and to all members of the Clinical Bioinformatics laboratory for constructive feedback.

## References

1. Caron B, Luo Y, Rausell A. NCBoost classifies pathogenic non-coding variants in Mendelian diseases through supervised learning on purifying selection signals in humans. Genome Biol 2019;20:32.

2. Chen S, Francioli LC, Goodrich JK et al. A genomic mutational constraint map using variation in 76,156 human genomes. Nature 2024;625:92–100.

3. Cheng J, Nguyen TYD, Cygan KJ et al. MMSplice: modular modeling improves the predictions of genetic variant effects on splicing. Genome Biol 2019;20:48.

4. Ellingford, Ahn, Bagnall et al., Recommendations for clinical interpretation of variants found in non-coding regions of the genome, Genome Medicine (2022) 14:73 10.1186/s13073-022-01073-3

5. Fu Y, Liu Z, Lou S et al. FunSeq2: a framework for prioritizing noncoding regulatory variants in cancer. Genome Biol 2014;15:480.

6. Ionita-Laza I, McCallum K, Xu B et al. A spectral approach integrating functional genomic annotations for coding and noncoding variants. Nat Genet 2016;48:214–20.

7. Jaganathan K, Kyriazopoulou Panagiotopoulou S, McRae JF et al. Predicting Splicing from Primary Sequence with Deep Learning. Cell 2019;176:535-548.e24.

8. Karczewski KJ, Francioli LC, Tiao G et al. The mutational constraint spectrum quantified from variation in 141,456 humans. Nature 2020;581:434–43.

9. Kaur G, Perteghella T, Carbonell-Sala S et al. GENCODE: massively expanding the lncRNA catalog through capture long-read RNA sequencing. 2024, DOI: 10.1101/2024.10.29.620654.

10. Kernohan KD, Boycott KM. The expanding diagnostic toolbox for rare genetic diseases. Nat Rev Genet 2024;25:401–15.

11. Kircher M, Witten DM, Jain P et al. A general framework for estimating the relative pathogenicity of human genetic variants. Nat Genet 2014;46:310–5.

12. Landrum MJ, Lee JM, Riley GR et al. ClinVar: public archive of relationships among sequence variation and human phenotype. Nucleic Acids Res 2014;42:D980–5.

13. OMIM - (OMIM.ORG). https://www.omim.org/ (July 2, 2025, xdate last accessed) PFMG2025 contributors. PFMG2025-integrating genomic medicine into the national healthcare system in France. Lancet Reg Health Eur 2025, doi: 10.1016/j.lanepe.2024.101183.

14. Sherry ST. dbSNP: the NCBI database of genetic variation. Nucleic Acids Res 2001;29:308–11.

15. Smedley D, Schubach M, Jacobsen JOB et al. A Whole-Genome Analysis Framework for Effective Identification of Pathogenic Regulatory Variants in Mendelian Disease. Am J Hum Genet 2016;99:595–606.

16. Schubach M, Maass T, Nazaretyan L et al. CADD v1.7: using protein language models, regulatory CNNs and other nucleotide-level scores to improve genome-wide variant predictions. Nucleic Acids Res 2024;52:D1143–54.

17. Schubach M, Nazaretyan L, Kircher M. The Regulatory Mendelian Mutation score for GRCh38. GigaScience 2022;12, DOI: 10.1093/gigascience/giad024.

18. Stenson PD, Mort M, Ball EV et al. The Human Gene Mutation Database (HGMD®): optimizing its use in a clinical diagnostic or research setting. Hum Genet 2020;139:1197–207.

19. Wagner N, Çelik MH, Hölzlwimmer FR et al. Aberrant splicing prediction across human tissues. Nat Genet 2023;55:861–70.

20. Walker LC, De La Hoya M, Wiggins GAR et al. Application of the ACMG/AMP framework to capture evidence relevant to predicted and observed impact on splicing; recommendations from the CLINGEN SVI splicing subgroup. 2023, DOI: 10.1101/2023.02.24.23286431.

21. Zhou J, Troyanskaya OG. Predicting effects of noncoding variants with deep learning–based sequence model. Nat Methods 2015;12:931–4.

22. Zoonomia Consortium, Genereux DP, Serres A et al. A comparative genomics multitool for scientific discovery and conservation. Nature 2020;587:240–5.

