## Supplementary Material for "NCBoost v2: a classifier for non-coding variants in Mendelian diseases"

### Supplementary Text

#### Methods

**High-confidence set of pathogenic variants.**

Two sets of high confidence pathogenic variants were aggregated: i) regulatory disease-causing mutations (DM) from the Human Gene Mutation Database, HGMD professional version (Stenson *et al.* 2020), accessed on the 01/04/2024, and ii) Single Nucleotide Variants (SNVs) annotated as "pathogenic" and without conflicting assertions from ClinVar database (Landrum *et al.* 2014), https://ftp.ncbi.nlm.nih.gov/pub/clinvar/, accessed on the 23/03/2024.

**Mapping and annotation of non-coding SNVs**

Only SNVs located on autosomes and gonosomes were considered throughout the study. Variants were associated with genes and genetic regions using the gene-based function of the ANNOVAR software (version 2020-06-07, (Wang, Li and Hakonarson 2010)) based on the CRCh38 RefSeq assembly version. Variants annotated as exonic, or within 10 base pairs (bp) from a splice site (ANNOVAR *splicing_threshold*=10) of a protein-coding gene, or associated with non-coding RNAs, were filtered out. Variants associated with several regions and/or several genes were handled according to the following criteria: i) the default precedence rule for gene-based annotation of ANNOVAR was applied to prioritize a region annotation over others: exonic = splicing > ncRNA > UTR5 = UTR3 > intronic > upstream = downstream > intergenic; ii) for variants mapping to equivalent priority regions but to different genes, the variant's closest protein-coding gene was retained, as defined by the shortest sequence distance between the variant and either the transcription start site (TSS) or transcription end site (TES) of flanking genes; and iii) variants located at the same distance from each flanking genes were labelled as "conflicting" and removed from further analyses. The final set of non-coding variants was constituted of SNVs associated with protein-coding genes and annotated as intronic, 5'UTR, 3'UTR, upstream, downstream, or intergenic variants. Upstream and downstream variants were defined as those closer than 1 kb from the transcription start site (TSS) and the transcription end site (TSE), respectively.

**Curation of pathogenic non-coding variants causing monogenic Mendelian diseases**

Further curation steps were applied to the aggregated set of pathogenic non-coding variants associated with protein-coding genes (**Figure S1**): (i) Variants with conflicting gene associations between the source database and the previously described ANNOVAR gene-based annotation were filtered out. (ii) To further refine the quality of the pathogenic variant set, we assessed the underlying genetic architecture of their associated gene as reported by OMIM database (OMIM). As in the first release of NCBoost, we followed similar filtering steps as detailed in Chong et al

(Chong *et al.* 2015): OMIM data files were downloaded from https://www.omim.org/downloads/ on the 12/11/2024. Genes were mapped to their corresponding phenotypes, and the phenotypic descriptions were used to assess the corresponding genetic architecture of the phenotype: i) phenotypic description containing the keyword "somatic" were flagged as somatic, those containing the keywords "risk", "quantitative trait locus", "QTL", "{", "[" or "susceptibility to" were flagged as "complex". We labelled as Mendelian disease genes those with a Clinvar supporting evidence level of 3 (i.e. disease molecular basis is known) and not flagged as somatic. These genes were further split in between monogenic mendelian disease genes (MMDGs, n= 4564) and complex mendelian disease genes (CMDGs, n=611) based on their "complex" flag. Among those, n=283 genes were associated with both monogenic and complex mendelian diseases. In addition, we further inspected whether the pathogenic non-coding SNVs were found as homozygous or heterozygous in GnomAD (Chen *et al.* 2024) (version 4.1) in both exome and genome data, and variants reported in homozygosis in at least one carrier were filtered out. After the different filtering steps, only 2336 high-confidence SNVs mapping to intronic, UTR5, UTR3, upstream, downstream and intergenic regions of protein-coding genes, causing monogenic mendelian diseases and not found as homozygous in GnomAD were retained for the purpose of this study.

**Common variants without known clinical consequences**

Variants from the general population were downloaded from dbsnp (Sherry 2001) (https://www.ncbi.nlm.nih.gov/variation/docs/human_variation_vcf/, filename GCF_000001405.40) on the 28/01/2025, including all SNVs from build 156. Common SNVs were extracted based on the annotation provided by dbsnp, corresponding to an allele frequency threshold of 0.01 (i.e. 1%). Common variants were first mapped to non-coding regions and cis-proximal genes using ANNOVAR gene-based annotation, and processed as described previously. After such filtering steps, a set of 16,169,467 common non-coding variants associated with protein-coding genes were retained for further analyses.

**State-of-the-art scores for the classification of pathogenic non-coding SNVs**

Pre-computed scores for two non-coding variant prioritization methods were downloaded for benchmarking purposes: CADD (version 1.7) (Schubach *et al.* 2024) and ReMM (for GRCh38 genome assembly) (Schubach, Nazaretyan and Kircher 2022).

**Feature extraction for non-coding SNVs**

The features used in this study are recapitulated in **Table S1** and are separated in the following four main categories:

**A - Long-term interspecies sequence conservation features**

To assess long-term sequence conservation, several scores assessing non-neutral mutation rates based on multiple non-human species alignment were extracted from CADD v1.7: i) the Genomic Evolutionary Rate Profiling (Gerp) (Davydov *et al.* 2010) neutral evolution score (GerpN), rejected substitution score (GerpS), element score (GerpRS) and element score pvalue (GerpRSpvalue); ii) PhCons (Siepel *et al.* 2005) and PhyloP (Pollard *et al.* 2010) scores from three multi-species alignment (primates, mammals and vertebrates, excluding humans); iii) PhyloP scores computed on 43 genomes of primates from the Zoonomia project (Christmas *et al.* 2023) (ZooPriPhyloP); PhyloP scores computed on 241 genomes of vertebrates from the Zoonomia project (ZooVerPhyloP); Ultra Conserved Elements (UCE) and Runs of Contiguous Constraint (RoCC) scores as assessed by the Zoonomia project.

**B - Recent and ongoing selection in humans**

The background selection score (Bstatistic) (McVicker *et al.* 2009) was extracted from CADD v1.7. The Context Dependent Tolerance Score (CDTS) hg19 (Di Iulio *et al.* 2018) was downloaded and genomic positions were lifted to hg38 assembly using liftover. The adjusted Roulette mutation rate estimates (Roulette-AR) (Seplyarskiy *et al.* 2023), were extracted from CADD v1.7. Mean minor allele frequencies (MAF) were computed on GnomAD v4.1 (Chen *et al.* 2024) genome data for all individuals as well as for individuals from 9 major sub-populations: Africans and African Americans (AFR), Amish (AMI), Admixed Americans (AMR), Ashkenazi Jewish (ASJ), East Asians (EAS), Finnish (FIN), Middle-Eastern (MID), non-Finnish Europeans (NFE) and South Asians (SAS). For each position in the genome and for each subpopulation, the mean MAF was calculated over the corresponding sub-population MAFs of all multiallelic loci located within +/- 500bp from the assessed position, excluding the MAF at the position.

**C - Gene conservation Features**

To assess the disease-causing potential of genes associated with non-coding variants, several gene-level conservation and mutation tolerance features were considered: the ratio between the number of non-synonymous variants and synonymous variants in primates (dn/ds) was downloaded from (Rausell *et al.* 2014); gene ages, estimated based on the presence or absence of ortholog genes in vertebrates, were downloaded from (Popadin *et al.* 2014). The probability of loss-of-function Intolerance score (pLI), the loss-of-function observed / expected upper bound fraction score (loeuf) and the deviation of observed counts from the expected number of synonymous (zscore_syn) and missense (zscore_mis) variants computed on GnomAD v4.1 individuals were downloaded from https://gnomad.broadinstitute.org/data#v4-constraint (Chen *et al.* 2024). The Gene Damage Index (GDI) was obtained from (Itan *et al.* 2015), and the non-coding residual variation intolerance score (ncRVIS), non-coding GERP (ncGERP), RVIS genome-wide percentile score (RVIS_percentile) and protein-coding GERP (pcGERP) scores were obtained from (Petrovski *et al.* 2015). Gene symbols and HUGO Gene Nomenclature Committee gene ids (HGNC (Yates *et al.*)) were mapped to Ensembl stable IDs (ENSG) obtained from Ensembl Biomart (Dyer *et al.* 2025) (human genome assembly version CRCh38.p14, accessed on the 24/04/2024) through its python API (pybiomart 0.2.0). When gene-level features were available for different isoforms of the same gene, the transcript with the highest amount of supporting evidence was selected, as annotated by Ensembl Biomart and Matched Annotation from NCBI and EMBL-EBI (MANE) (Morales *et al.* 2022) transcripts databases.

**D - Sequence and context features**

The percentage of CpG and CG in +/- 75bp around each position were extracted from CADD v1.7. SpliceAI (Jaganathan *et al.* 2019) precomputed scores were downloaded from https://github.com/Illumina/SpliceAI, release 1.3, and, following the author's recommendations, the maximum value among the 4 splice-altering predicted scores across all possible substitutions at each position was used to recapitulate the splice-altering potential of each position. In addition, the non-coding regions (i.e. intronic, UTR5, UTR3, upstream, downstream and intergenic) overlapping each position, as predicted by ANNOVAR, were one-hot-encoded as used as binary features in order to learn region-specific feature interactions.

**NCBoost v2 training framework and cross-training evaluation.**

NCBoost v2 training framework is similar to the first release, although it was implemented in python for better performance. NCBoost v2 training is based on the python implementation of eXtreme Gradient Boosting (XGBoost) v2.1.3 (https://xgboost.readthedocs.io/en/), with hyperparameters similar to the ones used in the first release of NCBoost (Caron, Luo and Rausell 2019), i.e.: eta = 0.01, gamma = 10 and max_depth = 25, in order to avoid overfitting. All the features used for training, recapitulated in **Table S1**, were used for training without prior standardization or normalization. To avoid cross-contamination between the training set and downstream/future application sets, NCBoost v2 was trained as a bundle of 10 models rather than a single model, in order to prevent training-testing cross contamination by design while reducing overfitting and ascertainment bias: First, the complete protein-coding gene list (N = 19,433 genes) was randomly split into 10 genomic partitions, equally stratified across chromosomes and each containing a similar amount of genes bearing and not bearing high-quality non-coding pathogenic variants curated for this study (see above). For the purpose of this work, each high-quality pathogenic non-coding SNVs associated with monogenic mendelian diseases (*i.e.* positive variants) were associated with 10 random common SNVs without clinical consequences (i.e. negative variants) mapped to the same non-coding region of protein coding genes from the same genomic partition. A maximum of 1 positive and 1 negative variants associated with the same gene were sampled. During model training, to prevent the over-representation of well characterized disease genes, a maximum of 1 pathogenic variant per gene was sampled, along its 10 associated region-matched common variants. In the case that, throughout such random sampling, there were not enough eligible common variants available fulfilling the above criteria, the corresponding pathogenic variant were filtered out. After the previous steps, a total of 687 pathogenic and their region-matched 6870 non-pathogenic variants were collectively used for model training. In summary, NCBoost v2 was trained as a bundle of 10 models, each being trained on the region-matched set of positive and negative variants from 9 of the 10 partitions, and being applied to score variants associated with genes from the held-out partition. Thus, each variant will receive one single score, attributed by the model whose training excluded any variant associated with the same gene. Variants from the training set scored in such a way were used for cross-training evaluation as reported in **Table 1**.

**Feature importance computation**

The relative importance of the features used for the training of NCBoost v2 were computed as the average of the total gain of each feature over the 10 models. The importance of all features sums up to 1. The standard deviation of the total gain of each feature was also computed.

**Score regional bias assessment**

The median score for each region of each gene attributed to pathogenic variants (N = 2336) and common variants (N = 16,169,467) was computed for NCBoost v2, CADD and ReMM. Detrimental regional biases were evaluated by assessing whether the distribution of median scores attributed to UTR-5' common variants (*i.e.* the distribution of scores attributed to common variants with the highest median) was higher than the distribution of median scores attributed to pathogenic variants across a given non-coding region using a one-sided Mann-Whitney-Wilcoxon test.

**Independent testing on a high-quality set of manually curated pathogenic non-coding SNVs**

The manually-curated set of non-coding disease-causing variants published along the first release of NCBoost was used to test NCBoost v2. For the purpose of this test, and in order to prevent cross-contamination between the testing set and the training set, NCBoost v2 was retrained, using only variants absent from the previous release. This set of pathogenic variants was converted to hg38 coordinates using the python implementation of liftover v1.3.1 (Perez *et al.* 2024), and the 694 non-coding variants successfully mapped to hg38 coordinates were further mapped to non-coding regions and genes by ANNOVAR and annotated as previously described. In addition, for each pathogenic variant, a region-matched set of random common variants were sampled from dbsnp as described above, following a 1 pathogenic to 10 common variants ratio. In the case that, throughout such random sampling, there were not enough eligible common variants available fulfilling the above criteria, the corresponding pathogenic variant were filtered out. After the previous steps, a total of 687 pathogenic and their region-matched 6870 non-pathogenic variants were used as an independent test set.

**Validation on a high-quality set of pathogenic non-coding variants**

Disease Mutations (DM) from the HGMD 2025.1 release were downloaded and annotated as previously described. A total of N = 211 DM non-coding variants, involving 158 protein-coding genes and absent from previous releases were obtained. As in the previous sets, for each pathogenic variant, a region-matched set of random common variants were sampled from dbsnp as described above, following a 1 pathogenic to 10 common variants ratio. In the case that, throughout such random sampling, there were not enough eligible common variants available fulfilling the above criteria, the corresponding pathogenic variant were filtered out. After the previous steps, a total of 151 pathogenic and their region-matched 1510 non-pathogenic variants were used as validation set.

**Simulation of disease genomes**

CRCh38 whole genome sequencing data from the 1000 Genome Project phase 3 (The 1000 Genomes Project Consortium *et al.* 2015) were downloaded from https://www.internationalgenome.org/data, and 100 genomes from European individuals were randomly sampled: HG00255, HG00237, HG00097, HG00154, HG00142, HG00252, HG00235, HG00100, HG00151, HG00134, HG00244, HG00130, HG00246, HG00126, HG00149, HG00152, HG00121, HG00112, HG00129, HG00231, HG00109, HG00114, HG00102, HG00122, HG00143, HG00123, HG00148, HG00238, HG00110, HG00135, HG00261, HG00136, HG00233, HG00099, HG00157, HG00137, HG00259, HG00256, HG00140, HG00243, HG00150, HG00111, HG00155, HG00145, HG00103, HG00262, HG00253, HG00260, HG04303, HG00264, HG00120, HG00116, HG00108, HG00240, HG01789, HG00234, HG00160, HG00251, HG00254, HG00245, HG00263, HG00242, HG00119, HG00127, HG00158, HG00139, HG00132, HG00096, HG00141, HG01334, HG04301, HG04302, HG00107, HG00117, HG01791, HG00101, HG00115, HG00257, HG00113, HG00133, HG00118, HG00131, HG00104, HG00258, HG00232, HG00156, HG00106, HG00138, HG00250, HG00239, HG00265, HG00146, HG01790, HG00249, HG00128, HG00159, HG00105, HG02215, HG00125 and HG00236. SNVs were mapped and annotated as described above, and only variants associated with non-coding regions (*i.e.* intronic, UTR5, UTR3, upstream, downstream and intergenic) of protein-coding genes were used for the analysis. To simulate disease genomes, we spiked in, within each genome, one of the 207 non-coding pathogenic variants from the validation set for which we could sample a random common variant mapping to the same non-coding region of the same gene. All variants were scored using NCBoost v2, CADD v1.7 and ReMM GRCh38, and the corresponding rank-percentiles were computed to ease comparison across scores. In case of ties, the minimum rank among the tied variants was attributed to all tied variants. Rank percentiles between spiked-in pathogenic and common variants and between rank-percentiles attributed to pathogenic variants by different methods were evaluated using one-sided Wilcoxon paired signed-rank tests (scipy 1.11.4).

**SpliceAI gene-level contextualization**

For each protein-coding gene, the top-scoring SpliceAI non-coding position was extracted and scored by NCBoost v2 and, for comparison purposes, the rank-percentiles were computed for both scores. In case of ties, the minimum rank was attributed to all tied entries. Next, for each score, we compared the distribution of rank-percentiles associated with non-disease genes (N = 12640) and MMDGs (N = 6128) using a two-sided Mann-Whitney-Wilcoxon test (scipy 1.11.4), as well as between MMDG rank-percentile distributions across scores using a two-sided Mann-Whitney-Wilcoxon rank-sum test and a two-sided Wilcoxon paired signed-rank test (scipy 1.11.4).

### Supplementary Figures

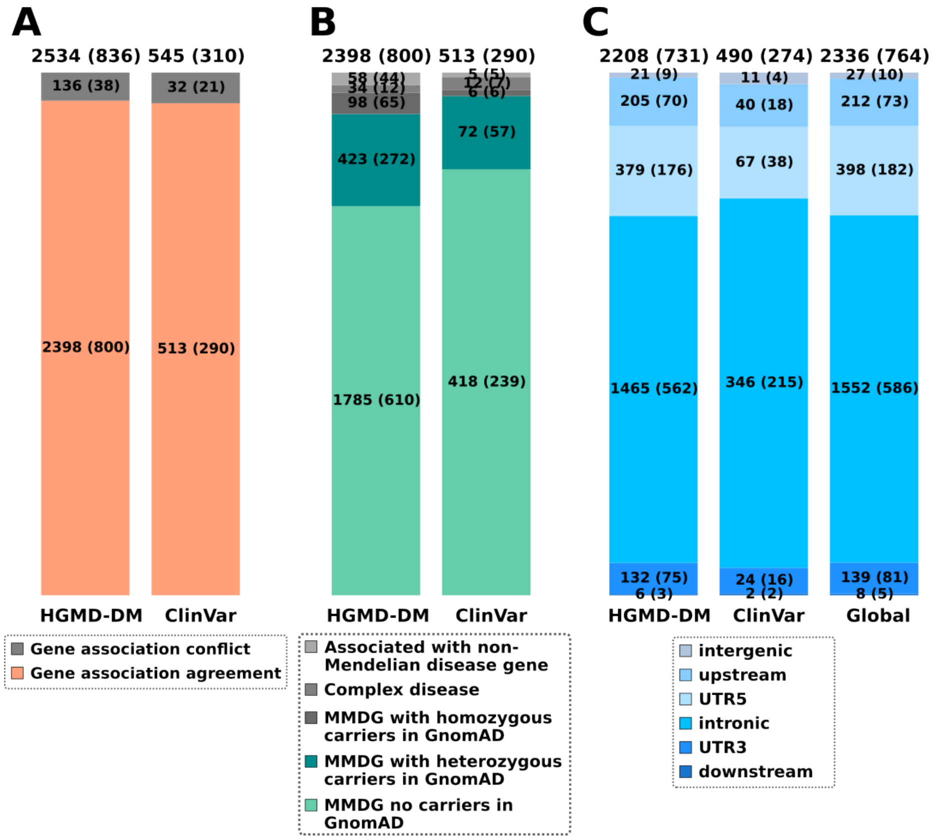


**Figure S1. Curation steps followed to obtain high-confidence pathogenic non-coding SNVs associated with monogenic Mendelian disease genes.** (A) Number of high-confidence pathogenic non-coding SNVs obtained from the Human Gene Mutation Database (HGMD-DM) and ClinVar, after filtering out exonic and splice-site SNVs, as well as SNVs associated with non-coding RNAs (Methods and Supplementary Text). SNVs associated with a different gene in the reference database than the one identified by the gene-mapping approach used in this work (shown in grey) were filtered out. (B) The SNVs retained in (A) were further categorized based on the gene they were associated with, according to their OMIM gene disease-type annotation: i.e., non-Mendelian disease, complex Mendelian disease, and monogenic Mendelian disease genes (MMDGs). Only SNVs associated with MMDGs and not observed in a homozygous state in any individual from gnomAD (shown in green) were retained for further analysis. (C) Non-coding region distribution of the final set of high-quality non-coding variants causing monogenic Mendelian diseases. For each region type, the corresponding number of variants is indicated, together with the number of unique genes involved (shown in parentheses). Total counts are reported above each barplot.

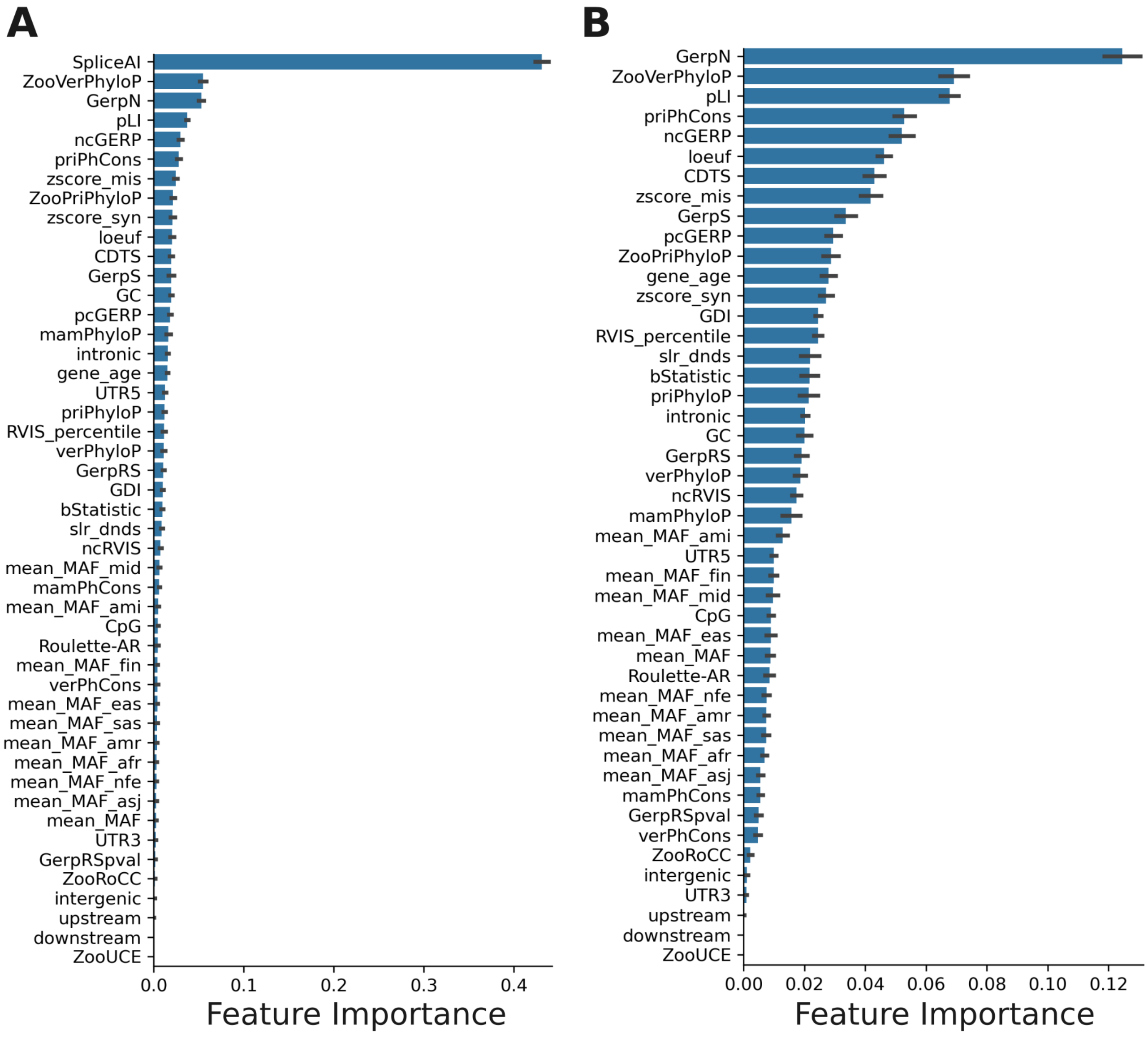


**Figure S2: Relative importance of NCBoost v2 features**

Figure shows the relative importance of the NCBoost v2 framework, including (**A**) or excluding (**B**) SpliceAI from the training features (**Methods** and **Supplementary Text**). Each barplot represents the mean importance over the 10 models, and the error bars represent the corresponding standard deviation. The x-axis of panels (A) and (B) are in different scales to ease visualization.

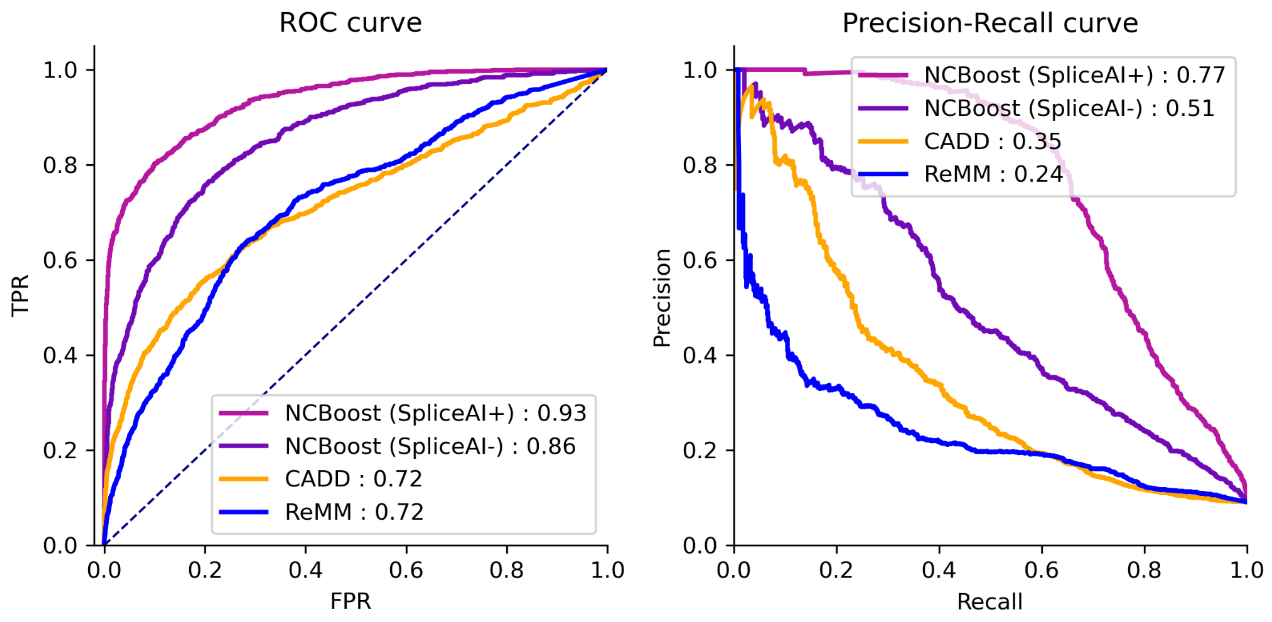


**Figure S3: Genomic-partition cross validation of NCBoost v2 against state-of-the-art methods**

Receiver operating characteristic and precision-recall curves of NCBoost v2, CADD v1.7 and ReMM, evaluated through genomic-partition cross validation on a set of N=763 high-quality pathogenic non-coding SNVs implicated in monogenic Mendelian diseases and N=7630 non-coding SNVs (**Methods** and **Supplementary Text**). The performance of NCBoost v2 is shown for a model trained with or without SpliceAI as a feature, indicated as NCBoost SpliceAI+ (purple) and NCBoost SpliceAI- (violet), respectively. The AUROC and AUPR values for each method are reported in the legends.

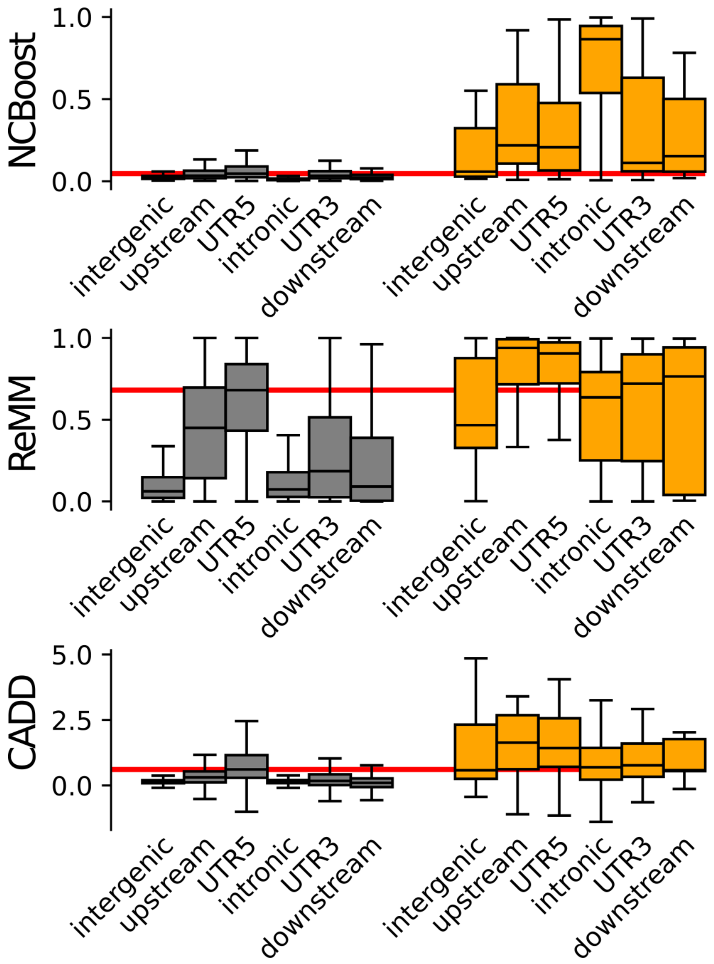


**Figure S4: Distribution of pathogenic scores of non-coding SNVs according to the type of the affected genomic region.**

Boxplots in the panels show the genome-wide distribution of per-region median scores of all non-coding SNVs associated with a given protein-coding gene (**Methods** and **Supplementary Text**). Two sets of non-coding SNVs are represented: n=16,169,467 common SNVs without clinical assertions (collectively associated with 18,196 protein-coding genes; in grey) and n=2334 high-confidence non-coding pathogenic variants (collectively associated with 763 monogenic Mendelian disease genes; in orange). Six types of genomic regions are depicted: intergenic, intronic, 3'UTR, 5'UTR, upstream and downstream regions of associated genes. Three scores are represented: NCBoost v2 (top) ReMM (middle); and CADD (bottom). The horizontal red line is depicted in each panel at the median value of the 5'UTR distribution for common non-coding SNVs. **Table S3** reports the one-sided Wilcoxon test p-values evaluating the null hypothesis that the median score distribution in 5'UTR for common non-coding SNVs is different than the distribution of the scores for pathogenic variants in the 6 types of genomic regions evaluated, i.e: intergenic, intronic, 3'UTR, 5'UTR, upstream and downstream.

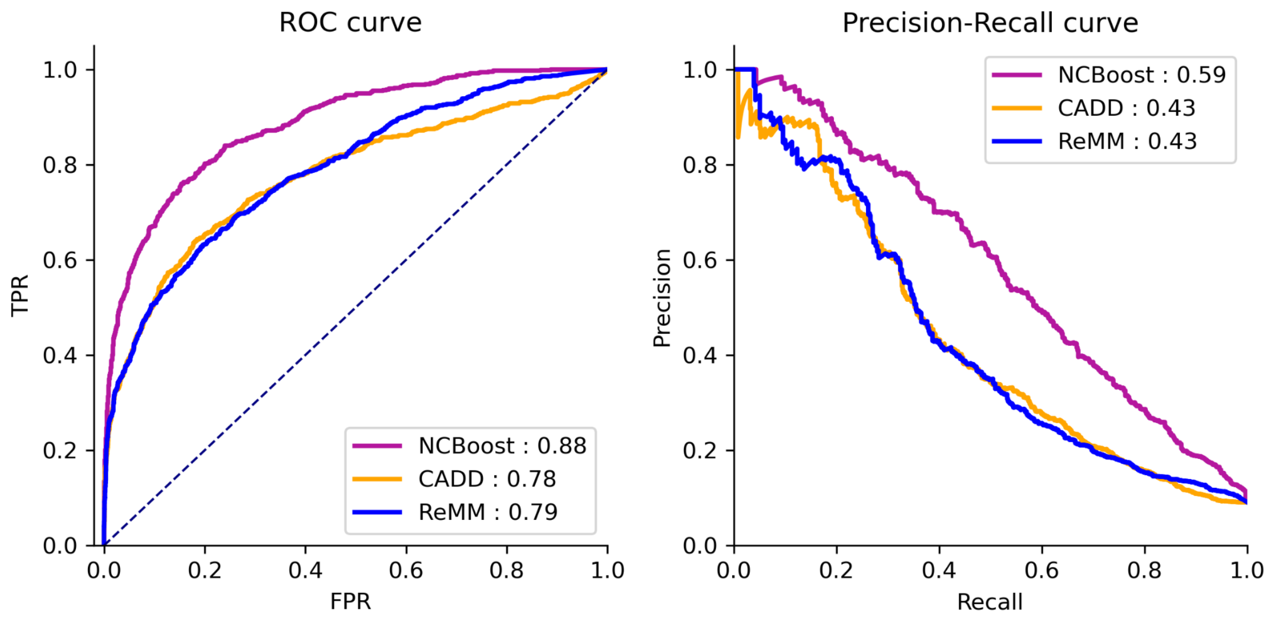


**Figure S5: Testing NCBoost v2 and alternative methods on a manually-curated set of non-coding pathogenic variants**

Receiver operating characteristic and precision-recall curves of NCBoost v2 (purple), CADD v1.7 (orange) and ReMM (blue), evaluated on a manually curated high-confidence set of N=687pathogenic non-coding SNVs implicated in monogenic Mendelian diseases and N=6870 non-coding SNVs (**Methods** and **Supplementary Text**). The AUROC and AUPR values for each method are reported in the legends.

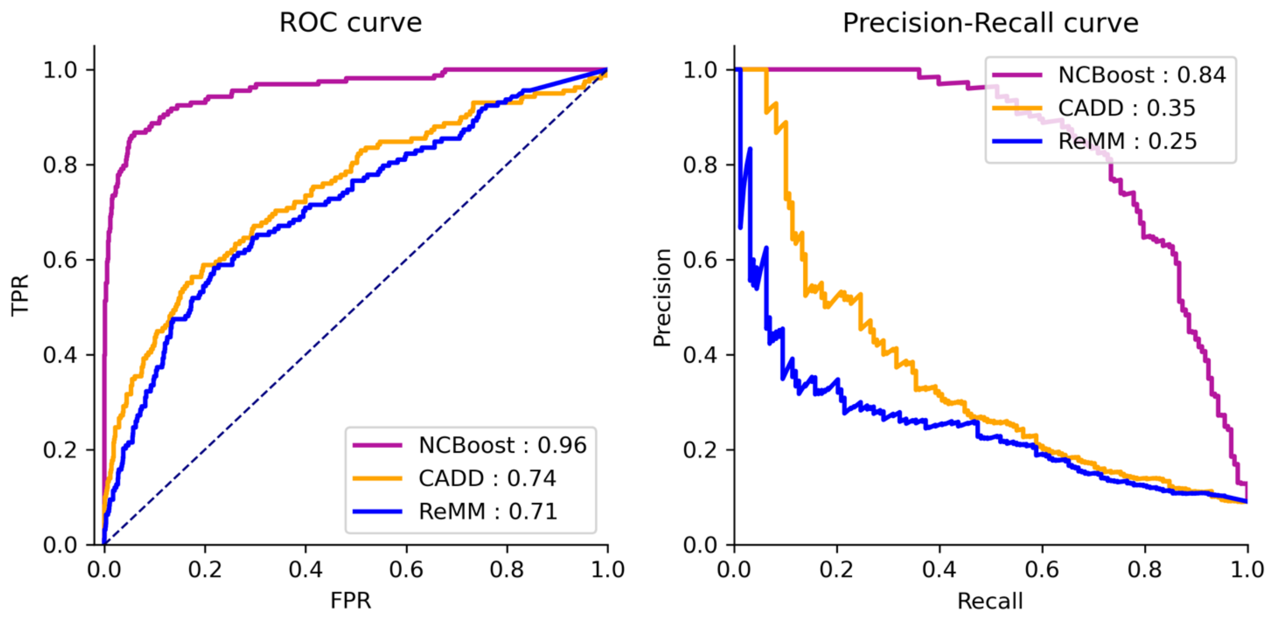


**Figure S6: Validation of NCBoost v2 and alternative methods on a recent high-confidence set of non-coding pathogenic variants**

Receiver operating characteristic and precision-recall curves of NCBoost v2 (purple), CADD v1.7 (orange) and ReMM (blue), evaluated on a recent high-confidence set of N=151 pathogenic non-coding SNVs implicated in monogenic Mendelian diseases and N=1551 non-coding SNVs (**Methods** and **Supplementary Text**). The AUROC and AUPR values for each method are reported in the legends.

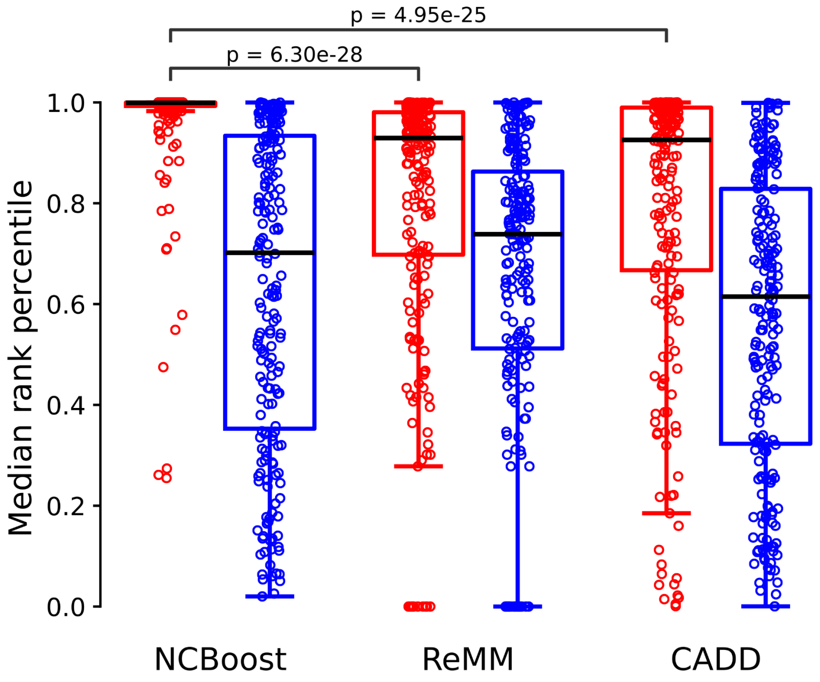


**Figure S7: Prioritization of disease-causing non-coding variants in individual genomes**

Median rank percentile (y-axis) across 100 simulated disease genomes of NCBoost v2, ReMM and CADD scores, attributed to 207 disease-causing non-coding variants (red dots) and the corresponding 207 region- and gene-matched common non-coding variants (blue dots, **Methods** and **Supplementary Text**). The boxplots show the overall distribution of the three different scores (x-axis), split over pathogenic (red) and common (blue) non-coding variants. NCBoost v2 provided the best within-individual ranking of non-coding pathogenic variants (median = 99.93%), significantly higher (one-sided paired Wilcoxon test p-value) than ReMM (median = 92.94, p-value = 4.95e-25) and CADD (median = 92.57%, p-value= 6.30e-28). The three scores ranked the non-coding pathogenic variants significantly higher than the common variants (one-sided paired Wilcoxon test): NCBoost v2 p-value = 1.41e-32 , ReMM p-value = 9.54e-11, and CADD p-value = 1.24e-13.

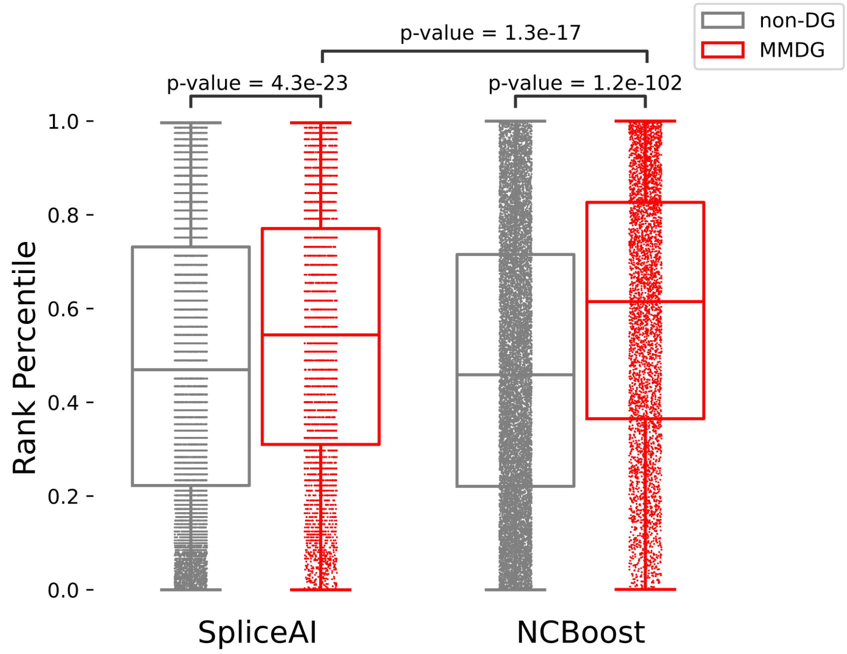


**Figure S8: NCBoost v2 improves SpliceAI detection of disease-causing splice-altering variants in monogenic Mendelian disease genes by providing gene-level contextualisation**

Boxplots show the rank-percentile distribution (y-axis) of SpliceAI (x-axis, left) and NCBoost v2 (x-axis, right) scores attributed to the top splice-altering position (as predicted by SpliceAI) of each non-disease (DG) protein-protein coding gene (grey) and monogenic Mendelian disease genes (MMDG, red). Each point represents the rank-percentile of the top splice-altering position of a given gene. Two-sided Mann-Whitney-Wilcoxon test p-values are shown over corresponding distributions.

### Supplementary Tables

**Table S1: Variant annotation features**

The table describes the 47 features used in NCBoost v2, including 15 new ones compared to the initial NCBoost version. Features are grouped in four categories: (A) long-term interspecies sequence conservation features; (B) recent and ongoing human sequence constraint features; (C) conservation features of the associated protein-coding gene, including both interspecies and in-human sequence conservation; and (D) sequence context features, such as GC and CpG content around the position, the type of overlapping non-coding region and the maximum SpliceAI score at the position. Further details are provided in **Supplementary Text**.

| Category | Sequence context | Evolutionary scale | Feature set | Feature abbreviation used in this work | Definition | Release of inclusion | References |
|---|---|---|---|---|---|---|---|
| Position-level and window-level features | Position-level | Interspecies (mammals) | A | GerpN | Genomic Evolutionary Rate Profiling (GERP++) neutral evolution score from sequence alignment in mammals, excluding humans | v1 | (Davydov *et al.* 2010) |
| | | Interspecies (mammals) | A | GerpS | GERP++ substitution rate from sequence alignment in mammals, excluding humans | v1 | (Davydov *et al.* 2010) |
| | | Interspecies (mammals) | A | GerpRS | GERP++ base-wise rejected substitution score in mammals, excluding humans | v2 | (Davydov *et al.* 2010) |
| | | Interspecies (mammals) | A | GerpRSpval | GERP++ base-wise rejected substitution pvalue in mammals, excluding humans | v2 | (Davydov *et al.* 2010) |
| | | Interspecies (vertebrates) | A | VerPhCons | Vertebrate PhastCons conservation score, excluding humans | v1 | (Siepel *et al.* 2005) |
| | | Interspecies (mammals) | A | mamPhCons | Mammalian PhastCons conservation scores, excluding humans | v1 | (Siepel *et al.* 2005) |
| | | Interspecies (primates) | A | priPhCons | Primate PhastCons conservation score, excluding humans | v1 | (Siepel *et al.* 2005) |
| | | Interspecies (vertebrates) | A | verPhyloP | Vertebrate PhyloP conservation score, excluding humans | v1 | (Pollard *et al.* 2010) |
| | | Interspecies (mammals) | A | mamPhyloP | Mammalian PhyloP conservation score, excluding | v1 | (Pollard *et al.* |

| | | | | | | | |
|---|---|---|---|---|---|---|---|
| | | | | | humans | | 2010) |
| | | Interspecies (primates) | A | priPhyloP | Primate PhyloP conservation score, excluding humans | v1 | (Pollard *et al.* 2010) |
| | | Interspecies (vertebrates) | A | ZooVerPhyloP | Zoonomia Vertebrate PhyloP conservation score from 241 vertebrate genomic alignments | v2 | (Christmas *et al.* 2023) |
| | | Interspecies (primates) | A | ZooPriPhyloP | Zoonomia Primate PhyloP conservation score from 43 primate genomic alignments | v2 | (Christmas *et al.* 2023) |
| | | Interspecies (mammals) | A | ZooRoCC | Zoonomia Runs of Contiguous Constraint for at least 20 contiguous bp. | v2 | (Christmas *et al.* 2023) |
| | | Interspecies (mammals) | A | ZooUCE | Zoonomia Ultra-Conserved Elements for 20 bp or longer segments with identical sequences in 98% of considered species | v2 | (Christmas *et al.* 2023) |
| | | Recent and ongoing in humans | B | Roulette-AR | Adjusted Roulette genome-wide mutation rate estimate | v2 | (Seplyarskiy *et al.* 2023) |
| | | Recent and ongoing in humans | B | bStatistic | Bstatistic: background selection score indicating the expected fraction of neutral diversity that is present at a site, based on human SNVs from Perlegen Sciences, HapMap phase II, the SeattleSNPs NHLBI Program for Genomic Applications, and the NIEHS Environmental Genome Project | v1 | (McVicker *et al.* 2009) |
| | 10-bp window | Recent and ongoing in humans | B | CDTS | The Context-Dependent Tolerance Score (CDTS) represents the difference between observed and expected variations in Humans. The expected variation is computed for each nucleotide genome-wide as the probability of variation of each nucleotide depending on its heptanucleotide context. CDTS was computed on 11,257 unrelated individuals. | v1 | (Di Iulio *et al.* 2018) |
| | 75-bp flanking region | N/A | D | GC | Percent GC in a window of ± 75 bp | v1 | (Harrison *et al.* 2024) |
| | | N/A | D | CpG | Percent CpG in a window of ± 75 bp | v1 | (Harrison *et al.* |

| | | | | | | | |
|---|---|---|---|---|---|---|---|
| | | | | | | | 2024) |
| | 500-bp flanking region | Recent and ongoing in humans | B | mean_MAF<br>mean_MAF_afr<br>mean_MAF_ami<br>mean_MAF_amr<br>mean_MAF_asj<br>mean_MAF_eas<br>mean_MAF_fin<br>mean_MAF_mid<br>mean_MAF_nfe<br>mean_MAF_sas | Mean minor allele frequency of variants in 1-kb window region calculated from GnomAD genome data (excluding the query position from the calculation), assessed on all individuals or on major subpopulations: Africans and African Americans (AFR), Amish (AMI), Admixed Americans (AMR), Ashkenazi Jewish (ASJ), East Asians (EAS), Finnish (FIN), Middle-Eastern (MID), non-Finnish Europeans (NFE) and South Asians (SAS). AMI, MID and SAS populations were added in NCBoost v2 | v1 & v2 | (Karczewski *et al.* 2020) |
| | 5000-bp flanking region | | D | SpliceAI | Splice-altering potential of the position, as predicted by SpliceAI | v2 | (Jaganathan *et al.* 2019) |
| Gene-level features | Coding region of the closest gene | Interspecies (primates) | C | dN/dS | Primate dN/dS ratio, providing a measure of the coding-sequence conservation across primates | v1 | (Rausell *et al.* 2014) |
| | | Recent and ongoing in humans | C | pLI | Probability of a gene to be intolerant to heterozygous and homozygous loss-of-function variants, assessed on GnomAD v4.1. | v1 | (Chen *et al.* 2024) |
| | | Recent and ongoing in humans | C | zscore syn<br>zscore mis | Deviation of observed counts from the expected number of synonymous (zscore_syn) and missense (zscore_mis) variants, assessed on GnomAD v4.1. | v2 | (Chen *et al.* 2024) |
| | | Recent and ongoing in humans | C | loeuf | The loss-of-function observed / expected upper bound fraction score (loeuf), assessed on GnomAD v4.1. | v2 | (Chen *et al.* 2024) |
| | | Recent and ongoing in humans | C | GDI | Gene Damage Index, a metric of the mutational damage that has accumulated in the general population, based on CADD scores and on the 1000 Genomes Project data | v1 | (Itan *et al.* 2015) |
| | | Recent and ongoing in humans | C | RVIS percentile | Residual Variation Intolerance Score (RVIS) percentile, measuring the departure from the average number of common | v1 | (Petrovski *et al. 2015*) |

| | | | | | | | |
|---|---|---|---|---|---|---|---|
| | | | | | functional mutations in genes with a similar amount of mutational burden in humans, assessed on 6503 whole exome from the NHLBI Exome Sequencing Project (ESP) | | |
| | | Interspecies (mammals) | C | pcGERP | Average GERP++ score of the protein-coding sequence of a gene (pcGERP) | v2 | (Petrovski *et al. 2015*) |
| | Non-coding region of the closest gene (a) | Recent and ongoing in humans | C | ncRVIS | RVIS score for promoter regions, 5’ UTR regions, and 3’ UTR regions of protein coding genes (ncRVIS) | v1 | (Petrovski *et al. 2015*) |
| | | Interspecies (mammals) | C | ncGERP | Average GERP++ score of the non-coding sequence of a gene (ncGERP) | v1 | (Petrovski *et al. 2015*) |
| | Phylo-genetic gene features | N/A | C | gene_age | Estimation of the origination time of genes from the presence or absence of orthologs in the vertebrate phylogeny. | v1 | (Popadin *et al.* 2014) |

*bp* base pairs, *GERP* Genomic Evolutionary Rate Profiling, *RS* Rejected Substitution, *N/A* not applicable

(a) Non-coding region of the closes gene defined in the original publication of ncRVIS and ncGERP as the collection of 5’UTR, 3’UTR, and an additional non-exonic 250 bp upstream of transcription start site (TSS)

**Table S2: Per-region SpliceAI contribution to NCBoost v2 performance**

The table reports the genomic-partition cross-validation of of two NCBoost frameworks: NCBoost trained with or without SpliceAI as a feature (indicated as SpliceAI+ and SpliceAI-, respectively). The area under the receiver operating characteristic (AUROC) curve and precision-recall (AUPR) are shown based on a total of 736 high-quality non-coding pathogenic variants and the corresponding region-matched set of 7,360 random common non-coding variants. In addition to the global values, AUROC and AUPR values are also shown for the different region types evaluated independently.

| | **NCBoost v2 (SpliceAI-)** | | **NCBoost v2 (SpliceAI+)** | |
|---|---|---|---|---|
| Region type | AUROC | AUPR | AUROC | AUPR |
| Global | 0.86 | 0.51 | 0.93 | 0.77 |
| Upstream | 0.87 | 0.54 | 0.88 | 0.55 |
| 5'UTR | 0.81 | 0.41 | 0.81 | 0.40 |
| Intronic | 0.87 | 0.53 | 0.97 | 0.89 |
| 3'UTR | 0.85 | 0.51 | 0.85 | 0.56 |
| Downstream | 0.84 | 0.65 | 0.87 | 0.58 |
| Intergenic | 0.77 | 0.48 | 0.77 | 0.48 |

**Table S3: Regional bias of pathogenic score distributions**

One-sided Wilcoxon test p-values (non-corrected for multiple testing) evaluating the null hypothesis that the per-gene median pathogenicity score distribution in 5'UTR for common non-coding SNVs is lower than the corresponding distribution for pathogenic variants in the 6 types of genomic regions evaluated in this study, i.e: intergenic intronic, 3'UTR, 5'UTR, upstream and downstream. Significant p-values after multiple testing correction (<8,33e-03) are highlighted in bold. The reported p-values are associated with the distributions represented in **Figure S4**. The distribution of variants and genes per region are reported in **Figure S1**. We note the low numbers for pathogenic non-coding variants in intergenic and downstream regions, which may compromise statistical significance when those regions are evaluated independently, due to low sample size.

| | **Intergenic** | **Upstream** | **5'UTR** | **Intronic** | **3'UTR** | **Downstream** |
|---|---|---|---|---|---|---|
| **NCBoost v2** | 1.91e-01 | **1.41e-23** | **3.42e-41** | **1.26e-254** | **4.08e-14** | **3.47e-03** |
| **ReMM** | 6.92e-01 | **5.90e-16** | **8.16e-28** | 0e-00 | 2.15e-01 | 3.26e-01 |
| **CADD** | 3.65e-01 | **1.40e-11** | **1.52e-25** | 3.34e-02 | 4.22e-02 | 2.91e-01 |

#### References


Caron B, Luo Y, Rausell A. NCBoost classifies pathogenic non-coding variants in Mendelian diseases through supervised learning on purifying selection signals in humans. *Genome Biol* 2019;**20**:32.

Chen S, Francioli LC, Goodrich JK *et al.* A genomic mutational constraint map using variation in 76,156 human genomes. *Nature* 2024;**625**:92–100.

Chong JX, Buckingham KJ, Jhangiani SN *et al.* The Genetic Basis of Mendelian Phenotypes: Discoveries, Challenges, and Opportunities. *Am J Hum Genet* 2015;**97**:199–215.

Christmas MJ, Kaplow IM, Genereux DP *et al.* Evolutionary constraint and innovation across hundreds of placental mammals. *Science* 2023;**380**, DOI: 10.1126/science.abn3943.

Davydov EV, Goode DL, Sirota M *et al.* Identifying a High Fraction of the Human Genome to be under Selective Constraint Using GERP++. Wasserman WW (ed.). *PLoS Comput Biol* 2010;**6**:e1001025.

Di Iulio J, Bartha I, Wong EHM *et al.* The human noncoding genome defined by genetic diversity. *Nat Genet* 2018;**50**:333–7.

Dyer SC, Austine-Orimoloye O, Azov AG *et al.* Ensembl 2025. *Nucleic Acids Res* 2025;**53**:D948–57.

Harrison PW, Amode MR, Austine-Orimoloye O *et al.* Ensembl 2024. *Nucleic Acids Res* 2024;**52**:D891–9.

Itan Y, Shang L, Boisson B *et al.* The human gene damage index as a gene-level approach to prioritizing exome variants. *Proc Natl Acad Sci* 2015;**112**:13615–20.

Jaganathan K, Kyriazopoulou Panagiotopoulou S, McRae JF *et al.* Predicting Splicing from Primary Sequence with Deep Learning. *Cell* 2019;**176**:535-548.e24.

Karczewski KJ, Francioli LC, Tiao G *et al.* The mutational constraint spectrum quantified from variation in 141,456 humans. *Nature* 2020;**581**:434–43.

Landrum MJ, Lee JM, Riley GR *et al.* ClinVar: public archive of relationships among sequence variation and human phenotype. *Nucleic Acids Res* 2014;**42**:D980–5.

McVicker G, Gordon D, Davis C *et al.* Widespread Genomic Signatures of Natural Selection in Hominid Evolution. Nachman MW (ed.). *PLoS Genet* 2009;**5**:e1000471.

Morales J, Pujar S, Loveland JE *et al.* A joint NCBI and EMBL-EBI transcript set for clinical genomics and research. *Nature* 2022;**604**:310–5.

OMIM. Online Mendelian Inheritance in Man, OMIM®. World Wide Web URL: https://omim.org/.

Perez G, Barber GP, Benet-Pages A *et al.* The UCSC Genome Browser database: 2025 update. *Nucleic Acids Res* 2024;**53**:D1243–9.

Petrovski S, Gussow AB, Wang Q *et al. The Intolerance of Regulatory Sequence to Genetic Variation Predicts Gene Dosage Sensitivity. PLOS Genetics* 2015, https://journals.plos.org/plosgenetics/article?id=10.1371/journal.pgen.1005492

Pollard KS, Hubisz MJ, Rosenbloom KR *et al.* Detection of nonneutral substitution rates on mammalian phylogenies. *Genome Res* 2010;**20**:110–21.

Popadin KY, Gutierrez-Arcelus M, Lappalainen T *et al.* Gene Age Predicts the Strength of Purifying Selection Acting on Gene Expression Variation in Humans. *Am J Hum Genet* 2014;**95**:660–74.

Rausell A, Mohammadi P, McLaren PJ *et al.* Analysis of Stop-Gain and Frameshift Variants in Human Innate Immunity Genes. Quintana-Murci L (ed.). *PLoS Comput Biol* 2014;**10**:e1003757.

Schubach M, Maass T, Nazaretyan L *et al.* CADD v1.7: using protein language models, regulatory CNNs and other nucleotide-level scores to improve genome-wide variant predictions. *Nucleic Acids Res* 2024;**52**:D1143–54.

Schubach M, Nazaretyan L, Kircher M. The Regulatory Mendelian Mutation score for GRCh38. *GigaScience* 2023;**12**, DOI: 10.1093/gigascience/giad024.

Seplyarskiy V, Koch EM, Lee DJ *et al.* A mutation rate model at the basepair resolution identifies the mutagenic effect of polymerase III transcription. *Nat Genet* 2023;**55**:2235–42.

Sherry ST. dbSNP: the NCBI database of genetic variation. *Nucleic Acids Res* 2001;**29**:308–11.

Siepel A, Bejerano G, Pedersen JS *et al.* Evolutionarily conserved elements in vertebrate, insect, worm, and yeast genomes. *Genome Res* 2005;**15**:1034–50.

Stenson PD, Mort M, Ball EV *et al.* The Human Gene Mutation Database (HGMD®): optimizing its use in a clinical diagnostic or research setting. *Hum Genet* 2020;**139**:1197–207.

The 1000 Genomes Project Consortium, Corresponding authors, Auton A *et al.* A global reference for human genetic variation. *Nature* 2015;**526**:68–74.

Wang K, Li M, Hakonarson H. ANNOVAR: functional annotation of genetic variants from high-throughput sequencing data. *Nucleic Acids Res* 2010;**38**:e164.

Yates B, Braschi B, Gray KA *et al. Genenames.Org: The HGNC and VGNC Resources in 2017 | Nucleic Acids Research | Oxford Academic*. https://academic.oup.com/nar/article/45/D1/D619/2333908 (July 25, 2025, date last accessed)
